# The Impact of Flood Adaptation Measures on Affected Population’s Mental Health: A mixed method Scoping Review

**DOI:** 10.1101/2023.04.27.23289166

**Authors:** Fatima El-Mousawi, Ariel Mundo Ortiz, Rawda Berkat, Bouchra Nasri

## Abstract

The frequency and severity of floods has increased in different regions of the world due to climate change. Although the impact of floods on human health has been extensively studied, the increase in the segments of the population that are likely to be impacted by floods in the future makes it necessary to examine how adaptation measures impact the mental health of individuals affected by these natural disasters. The goal of this scoping review is to document the existing studies on flood adaptation measures and their impact on the mental health of affected populations, in order to identify the best preventive strategies as well as limitations that deserve further exploration. This study employed the methodology of the PRISMA-ScR extension for scoping reviews to systematically search the databases Medline and Web of Science to identify studies that examined the impact of adaptation measures on the mental health of flood victims. The database queries resulted in a total of 857 records from both databases. Following two rounds of screening, 9 studies were included for full-text analysis. Most of the analyzed studies sought to identify the factors that drive resilience in flood victims, particularly in the context of social capital (6 studies), whereas the remaining studies analyzed the impact of external interventions on the mental health of flood victims, either from preventive or post-disaster measures (3 studies). There is a very limited number of studies that analyze the impact of adaptation measures on the mental health of populations and individuals affected by floods, which complicates the generalizability of their findings. There is a need for public health policies and guidelines for the development of flood adaptation measures that adequately consider a social component that can be used to support the mental health of flood victims.

## 1. Introduction

Floods are broadly defined as overflowing water bodies such as rivers, streams, and main channels leading to inundations (Şen, 2018). These events are among the most common types of natural disasters, and the leading cause of natural disaster fatalities worldwide with an economic impact estimated in the range of billions of dollars (Doocy et al., 2013; Svetlana et al., 2015). Flood occurrence increased during the 20^th^ century due to climate change (Milly et al., 2002), and it is expected that the number of regions threatened by flood hazards will expand in the near future.

The continued threat that floods represent has motivated the study of their impact on human health. In this regard, it has been shown that flooding can greatly affect human populations. For example, it is estimated that mortality rates can increase by 50% following a flood event, with a concomitant increase in the risk of disease outbreaks such as hepatitis E, gastrointestinal diseases and leptospirosis (Alderman et al., 2012), as well increased rates of woundings, poisonings, and infections (Saulnier et al., 2017). Moreover, other studies have analyzed how floods affect mental health, showing that post-traumatic stress disorder, depression, and anxiety are among the consequences of this type of disaster (Mason et al., 2010).

On the other hand, it is known that adaptation measures, which are broadly defined as preventive interventions or interventions that respond to the effects of floods by adjusting, moderating, and coping with the risks (Allwood et al., 2014), can also impact human health. In particular, the different adaptation measures that exist in the case of floods, such as building barriers around properties, elevating appliances and furniture, installing pump systems, systematic evacuation and shelter preparation (Nofal and van de Lindt, 2020), usually involve broad geographical areas where large segments of the population live; therefore, the implementation of these measures is likely to have a direct effect not only on the physical health but also on the mental health of those affected by them. However, in contrast with the amount of literature that explores the impact of flood adaptation measures on physical health, the impact of adaptation measures on mental health remains relatively unexplored (Sharifi et al., 2021).

### Research question

The goal of this scoping review is to document the existing body of research on flood adaptation measures and their impact on the mental health of affected individuals and populations, in order to identify best practices, limitations, and areas that deserve further exploration in light of the expected increase of flood frequency and severity due to climate change.

## 2. Methods

The strategy used for this scoping review was developed and validated with the help of a librarian (Sylvie Fortin) from the Université de Montréal. The review was conducted following the recommendations of the PRISMA-ScR extension (Preferred Reporting Items for Systematic Reviews and Meta-Analysis: extension for Scoping Reviews (Tricco et al., 2018)). Three core concepts were included in the analysis: floods, mental health factors, and adaptation measures. For this scoping review, floods were defined as the overflowing of a stream or other body of water, or the accumulation of water over areas that are not normally submerged, including fluvial, flash, urban, pluvial, sewer, coastal, and glacial lake outburst floods (Field et al., 2012). Mental health was defined as the state of psychological and emotional well-being. Due to the relationship between mental health, coping skills (i.e. how individuals manage stressful situations), and self-esteem (Mann et al., 2004), these three factors were considered as indicators of mental health status. Finally, adaptation measures were defined as anticipatory, autonomous or planned actions that reduce the negative impact of floods while taking advantage of potential new opportunities, involving the adjustment of policies and actions because of observed or expected changes in climate (Richardson, 2010).

Finally, adaptation measures were defined as actions that aim at reducing the negative impact of floods, which can be taken by individuals, communities, or governments. At the individual level these include purchasing home insurance, warning or helping others, or adapting their household (Brink and Wamsler, 2019); at the community level, adaptation measures include planting trees in public spaces and preparing disaster response plans (Ebi and Semenza, 2008); at the government level, they can include the construction of flood protection systems or the adoption of renewable energy sources (Osberghaus et al., 2010).

### Eligibility Criteria

Inclusion and exclusion criteria were based on the Population, Intervention, Comparison, and Outcome (PICO) framework (Amir-Behghadami and Janati, 2020). The study population was defined as flood victims, the intervention was defined as flooding adaptation measures, and the outcome was defined as the mental health of the population. No specific study comparators were defined; hence, the comparator element of the PICO framework did not apply to this review. The criteria for exclusion were based on language, study setting, population, intervention, study design, and outcome. First and foremost, any record not written in English or French was excluded. As for the setting, any record about a pre or post-flood event caused by another natural disaster such as an earthquake, cyclone, or hurricane was excluded. Records referring to studies not conducted with flood victims, literature reviews, or non-study records were also excluded. Finally, any study whose outcomes were not mental health, or an indicator of mental health as defined above were also excluded. All records that met the criteria at the end of the screening process were included for full-text reading to assess whether they were relevant to this scoping review.

### Information Sources

Two databases were searched for this scoping review: Medline, which was accessed through the Ovid platform, and Web of Science. Medline was chosen as it is considered the reference library for health literature. Web of Science was used to obtain records on floods and flood adaptation measures from journals not indexed on Medline.

### Search Strategy

The search was first carried out on March 8th, 2021, and updated on September 26th, 2022 using the three core concepts of floods, mental health, and adaptation measures, as indicated before.

The following keywords were used for the first concept: *inundation, flood, heavy precipitation, rain, rains, river, lake, and high water*; for the second concept: mental health, psychological, substance abuse, mental disorder, mental illness and distress; and for the third concept keywords were *disaster, risk, reduction, preparedness, resilience, planning, management, adaptation, planning, strategy, measure, decision*, and *approach*. The search strategies for both databases are available in Supplementary Material.

### Selection of Sources of Evidence

All the queries were imported into Covidence, a systematic review software platform which automatically removed duplicate entries (https://www.covidence.org/). Covidence was used to perform the first round of screening. The review of titles and abstracts was done by two independent reviewers trained in global environmental health and in epidemiology (FEM and RB). In case of disagreement, a senior reviewer (BN) assessed the conflict and decided if the manuscript should be included for full-text review. In the second round of screening the same two reviewers (FEM, RB) read the full-text of each study and determined its inclusion based on the PICO framework and the exclusion criteria. In case of disagreement regarding the inclusion of the study for analysis, conflicts were solved by the senior reviewer (BN).

### Data Charting Process and Data Items

For data extraction, an Excel spreadsheet was used to obtain the following information from each study: authors, publication year, title, country, study design, year of the flooding event analyzed, sample size, data collection date, climate indicators, health indicators, methods stated in the abstract, results stated in abstract, detailed methods from the methods section, main findings from the results section, relevant comments, and paper citation. Data extraction was done by the same reviewers involved in the first round of screening.

### Synthesis of Results

The studies were grouped based on their focus to analyze mental health outcomes in communities or individuals affected by floods. For each study, information about the methodology and instruments used to collect data, sample sizes (when applicable), and principal findings is presented.

## 3. Results

### Selection of Sources of Evidence

The database queries resulted in 693 records obtained from Medline, and 266 records from Web of Science. Upon transfer of the records to the Covidence platform 102 duplicates were identified and removed, leading to a total of 857 records for title and abstract screening. After the first screening round, 18 studies that met the screening criteria were included for full-text reading. Following the second round of screening, 9 studies were relevant and included in the review (Figure 1).

**Figure 1.**
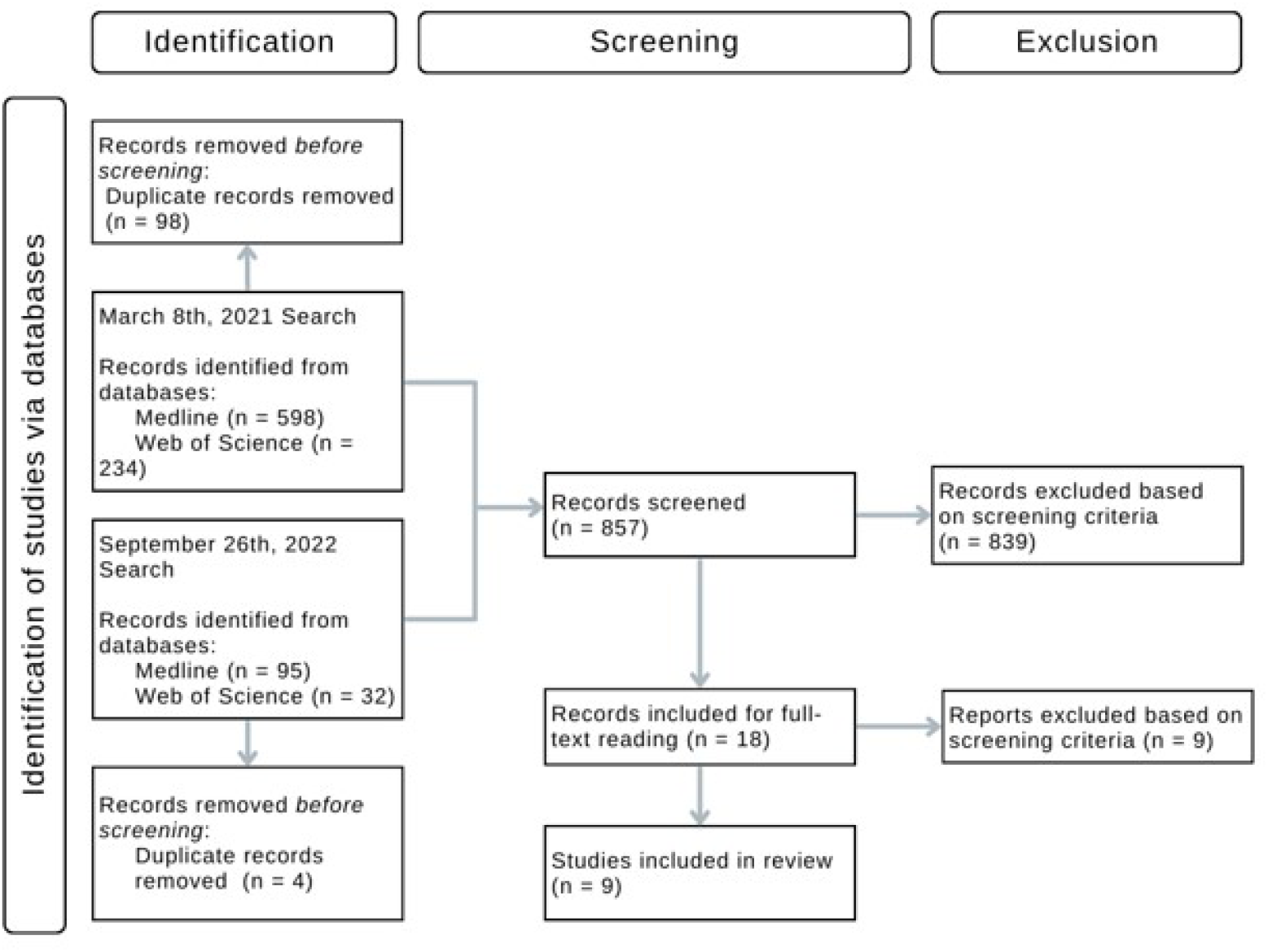
Flow diagram of the study selection process

### Characteristics of Included Studies

The included studies are presented in Table 1. Among them, five studies (56% of the total) used statistical models to determine the relationship between socio-economic covariates and mental health outcomes (Babcicky and Seebauer, 2017; Ludin et al., 2019; Masson et al., 2019; Song and Li, 2019; Zhong et al., 2020). On the other hand, four studies (44% of the total) used information collected via interviews surveys to qualitatively assess the impact of floods on the mental health of flood survivors (Brockie and Miller, 2017; Butler et al., 2018; Mustaffa et al., 2018; Shultz et al., 2013). Furthermore, two studies were conducted in China (Song and Li, 2019; Zhong et al., 2020), and two in Malaysia (Ludin et al., 2019; Mustaffa et al., 2018); the remaining studies were conducted in five different countries: Austria (Babcicky and Seebauer, 2017), Australia (Brockie and Miller, 2017), Germany (Masson et al., 2019), the United States of America (Shultz et al., 2013), and the United Kingdom (Butler et al., 2018).

**Table 1.**
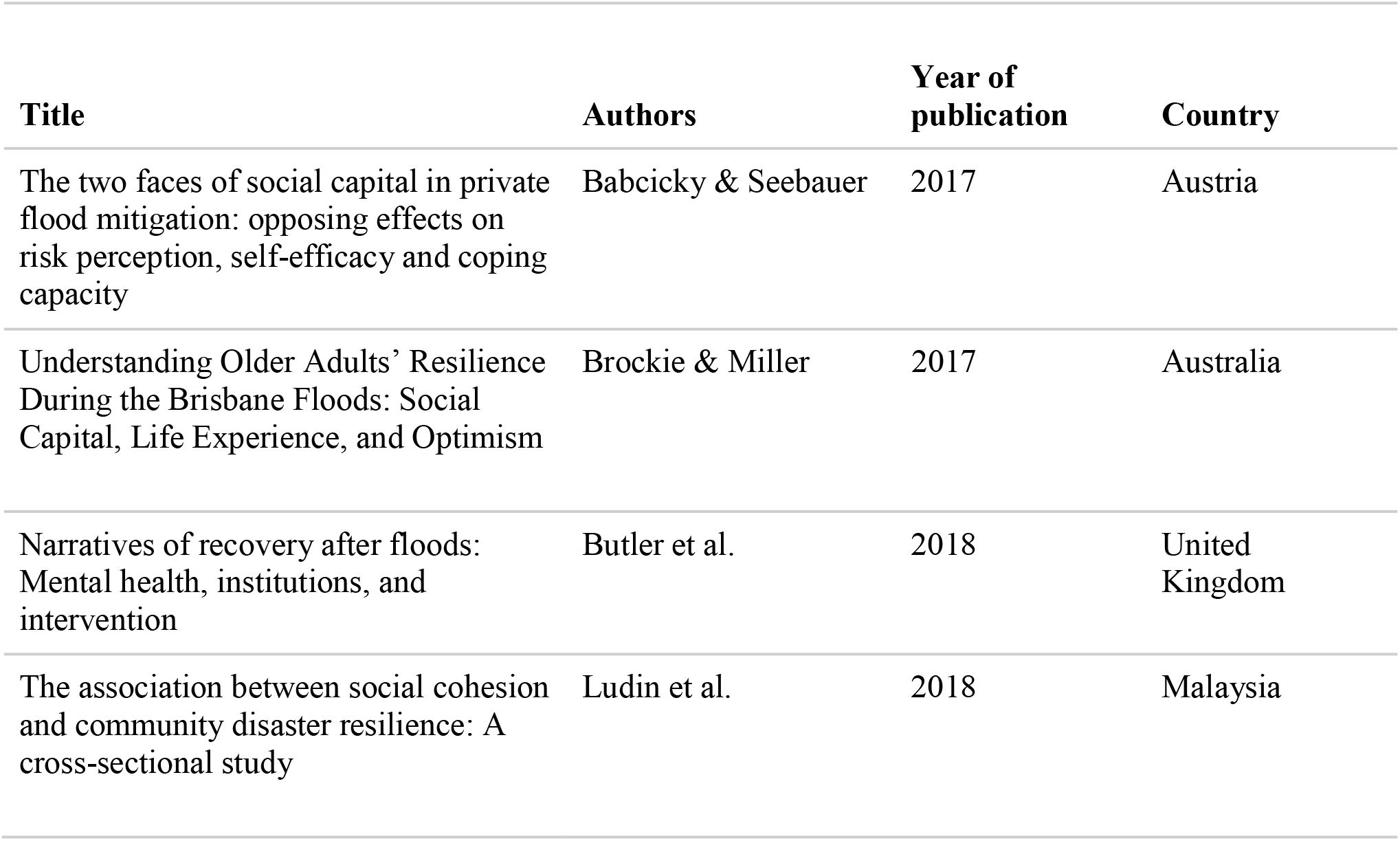
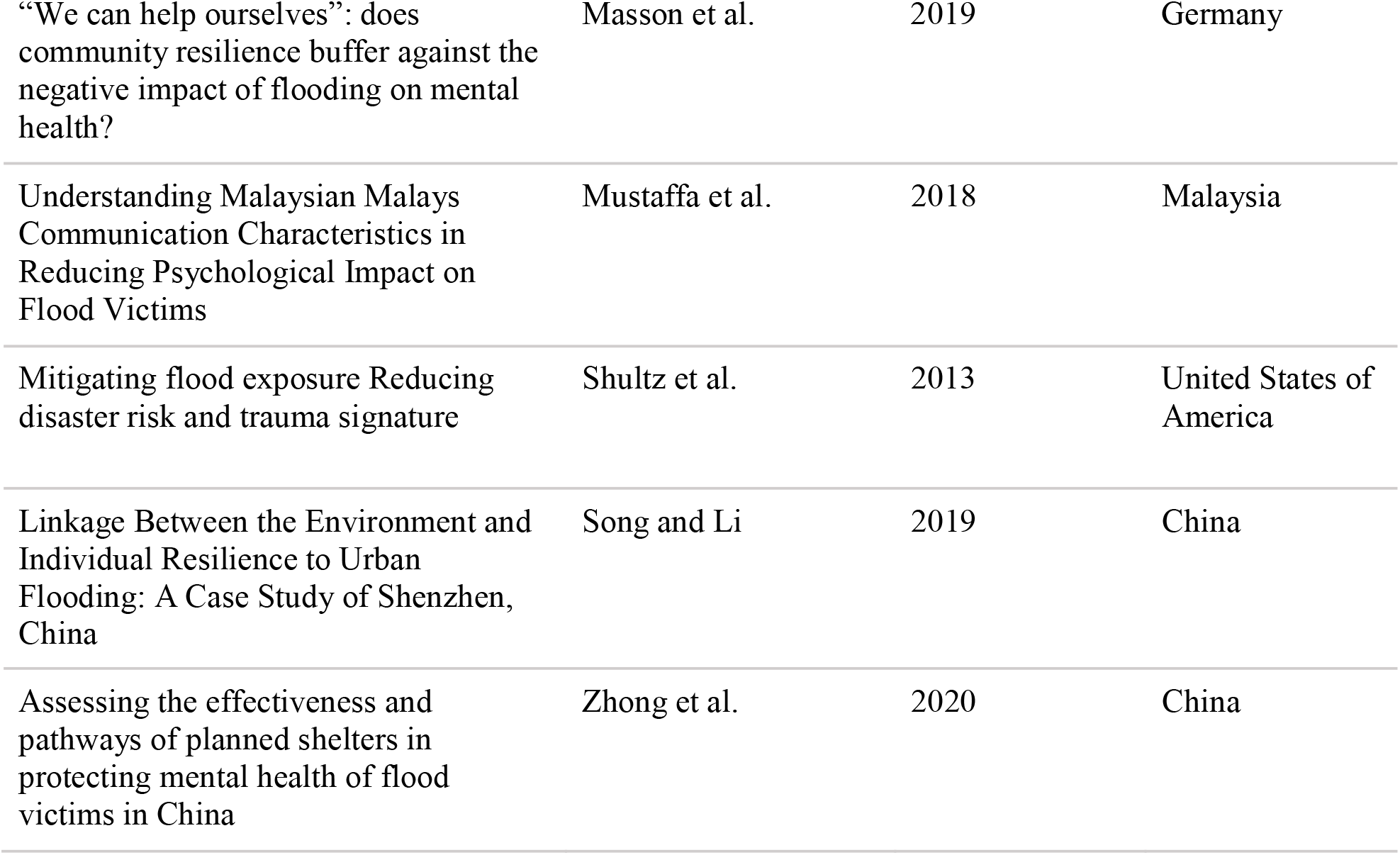
Articles included in the review.

### Results of Individual Sources of Evidence

Two categories of studies were identified based on their focus to analyze mental health outcomes in affected communities or individuals. The first category included studies that focused on resilience as an indicator of mental health, whereas the second category included studies that focused on the impact of external interventions on the mental health of flood victims, either from preventive measures (such as planned shelters) or post-disaster measures (such as evacuation).

### Summary of the included studies

#### Studies that focused on resilience as an indicator of mental health

Babcicky and Seebauer (2017) conducted a cross-sectional study where a Likert-scale survey was conducted in the cities of Oberwölz and Kössen in Austria, which experienced floods in 2011 and 2013, respectively. A total of 226 individuals from both cities were surveyed to obtain measured scores for cognitive risk perception, affective risk perception, combined (cognitive and affective) risk perception, self-efficacy, and cognitive social capital, besides socio-demographic characteristics. Multiple linear regression was conducted to identify the relationships between the variables. The study’s findings show that social capital increased perceived self-efficacy, contributed to coping capacity, decreased risk perception of private households (which were therefore less likely to apply flood mitigation measures), and boosted households’ belief in their own coping abilities. The authors conclude that informal social ties are essential for resilience against extreme flooding.

Brockie and Miller (2017) followed a qualitative approach by surveying a total of 10 participants aged 65 years and over who experienced flooding events in 2011 and 2013 while living in their own homes in the city of Ipswich in Queensland, Australia. After transcribing the interview verbatims, analysis was done following an inductive approach. The study found that social interactions, in the form of bonding capital from friends and family, were strongly linked to the practical and psychological resilience of older adults, besides their previous flood experience. The study also highlighted the change in the social capital network and communication avenues when comparing past flood experiences and recent ones.

Ludin et al. (2019) surveyed 386 respondents that experienced the floods of 2014 in Kelantan, Malaysia. The survey consisted of two parts: one that collected demographic information about the participants, and a second one that measured community cohesion and community resilience by using Buckner’s Index of Cohesion (which uses the answers to 18 items to measure neighborhood cohesion), and the Index of Perceived Community Resilience (which is obtained from a series of questions that address community resilience). The authors then estimated mean scores for BIC and IPCR and used these scores as the dependent variable in regression models that examined the influence of demographic factors in cohesion and resilience. This study showed that certain characteristics that indicate individual preparedness, such as participating in emergency teams, volunteering in activities, and having emergency disaster experience were associated with increased resilience and social cohesion.

Masson et al. (2019) surveyed 118 participants from the town of Simbach am Inn, in Germany, who experienced severe flash floods in 2016. The survey consisted of questions about the perceived consequences of the floods, flood-related psychological and physical distress, sense of coherence, life satisfaction, perceived collective social support, perceived interpersonal social support, and ego resilience, which respondents answered using a 6-point Likert scale which ranged from “not affected” to “very severe”. The association of the survey information and post-disaster mental health was analyzed using multiple hierarchical regression and path analysis, showing that perceived collective support (community resilience) was positively associated with post-disaster mental health and that low levels of community resilience and having a “very severe” perception of a flood event were associated with worse post-flood mental health outcomes.

Shultz et al. (2013) used trauma signature analysis (an evidence-based method that examines the interrelationship between population exposure to a disaster, and its inter-related physical and psychological consequences with the intent of providing timely, actionable guidance for effective mental health and psychosocial support(Shultz and Neria, 2013)) to examine the interrelationship between the populations of two demographically-comparable cities affected by floods in 2011 (Fargo and Minot, both located in North Dakota), and the physical and psychological consequences of the disaster. The authors used census and civic data to compare both cities, and established the physical hazard profile of the flood for each city based on government and local sources and expert consultation. Additionally, major disaster stressors, flood preparedness and flood response actions for each city were identified, and the trauma signature for each city was summarized by comparing the psychological risk factors for the flood events in each city. The authors found that because the city of Fargo applied flood prevention measures to reduce risk exposure and had strong resilience indicators, it was able to reduce the disaster risk and impact while diminishing the trauma signature in the population. On the other hand, the lack of prevention measures in the city of Minot resulted in the distress of the population as the city was inundated. The authors concluded that reducing risk exposure reduces flood trauma signature, both on the physical and psychological levels.

Finally, Song and Li (2019) surveyed 733 individuals from the city of Shenzhen in southeast China, a location that experienced multiple flooding events between 2012 and 2015, and collected socio-demographic factors of the participants and measures of disaster awareness (such as being confident in overcoming natural disasters, previous experience with natural disasters, and level of preparedness). The authors then used multiple hierarchical linear models to determine the association between socio-economic factors and the resilience of individuals that live in areas where floods occur, in order to determine coping and adaptive behaviors toward urban flooding (such as green spaces in and around the community and support from community organizations). The main findings of this study were that the perception of the living environment (such as the cleanliness of the physical environment and the presence of green spaces) is associated with increased individual resilience, which is also enhanced by having efficient and reliable social cohesion.

#### Studies that focused on the impact of external interventions on the mental health of flood victims

Zhong et al. (2020) analyzed the implementation of planned shelters to welcome internally displaced persons after floods in China. The study focused on individuals that experienced the flood event in June 2016 in the province of Anhui. For analysis, 338 flood victims from 69 planned shelters in the province of Anhui were considered as the intervention group, and 327 flood victims that did not live in shelters were considered as the control group. The study used multivariate logistic regression (using socio-economic information and flood exposure among other covariates) and structural equation modeling (using the effect of different policy interventions) to determine the impact of planned shelters on mental health. The findings of the study provided evidence that persons living in planned shelters had a lower prevalence of mental health problems than victims who remained in their own homes.

In the longitudinal study of Butler et al. (2018), two semi-structured qualitative interviews were conducted with 9 participants affected by floods that occurred in Somerset, in South West England (United Kingdom) in the winter of 2013, with a follow-up at 6 months and at 12 to 14 months. The study showed that, based on the narrative provided by the participants, indirect or direct actions taken by authorities and institutions in the face of floods (such as the process of evacuating the victims from their homes), as well as inaction, have implications for the mental health of the affected populations. Moreover, the authors indicated that community support alone is not enough, and hence, institutional support remains necessary, especially to address post-flood mental health issues.

Mustaffa et al. (2018) performed a qualitative study by conducting interviews with 13 respondents who were victims of the floods of 2014 in Malaysia. The verbatims of the interviews were coded using a comparative interpretive approach. The authors analyzed the answers of the participants using thematic data analysis techniques and following the guidelines for qualitative research of Creswell (Creswell and Poth, 2016) in order to identify the main factors discussed by the respondents. The authors reported that respect, sincerity, caring attitude, professionalism, and dignity maintenance were major values of Malaysian society that were reflected in the communication discourse of relief workers at disaster relief centers. The authors concluded that this type of language was required to reduce the psychological impact of the disaster on the victims and to ensure effective communication.

## 4. Discussion

The implementation of adaptation measures to manage floods should become more prevalent with the increase in the severity and frequency of these natural disasters. This scoping review collected studies from the literature on the impact of adaptation measures on the mental health of flood victims. Based on their analysis of mental health, the 9 studies that were included for full-text analysis (Table 1), were divided in two categories: 1) studies about resilience as an indicator of mental health, and 2) studies about other indicators of mental health in flood victims.

The first category included six studies that were focused on factors that influence resilience in flood victims. In a mental health context, resilience is broadly defined as the capacity to maintain mental health through an adversity and to positively adapt afterwards (Herrman et al., 2011; Saeed and Gargano, 2022). In the context of natural disasters and climate change, the ability to regenerate, reorganize, and redevelop to an improved state in the long term is also a component of resilience (Engle et al., 2014; Zhou et al., 2010). Considering that floods are expected to become more severe in the near future, there is a pressing need to develop resilience in individuals and communities that are likely to be affected by this type of disaster, in order to ensure that an improved state is indeed achieved.

In this regard, the studies of the first category identified various components of social capital as driving factors of resilience in flood victims (Table 2). In the context of natural disasters, social capital can be broadly defined as the social ties, and social networking of an individual that facilitates collective action (Jones and Clark, 2013; Smith et al., 2012). Specifically, the studies identified social ties (Babcicky and Seebauer, 2017), bonding (Brockie and Miller, 2017), community involvement (Ludin et al., 2019), social sources of support (Masson et al., 2019), community preparedness (Shultz et al., 2013), and social cohesion (Song and Li, 2019) as factors associated with resilience in individuals or communities affected by floods. These results provide a broad view of some of the factors that would need to be considered in the development of preemptive and post-flood intervention measures that are aimed at not only dealing with the physical aspect of this type of disaster, but that aim to ensure a positive mental health outcome for flood victims.

**Table 2.** Mental health outcomes assessed per included study.

| Study | Mental health outcomes |
| --- | --- |
| Babcicky & Seebauer | Informal social ties are essential factors for resilience against extreme flooding. This includes two distinct subcategories: cognitive and structural social capital. |
| Brockie & Miller | Three major themes that drove resilience in flood survivors were identified: social capital, previous experience of disasters, and sources of support. |
| Butler et al. | Institutional support has both positive and negative effects on the mental health of victims in a flood context. |
| Ludin et al. | Social cohesion increases flood victims' resilience and improves physical and mental health. |
| Masson et al. | Community resilience to flooding through collective social support buffers the negative mental health effects, including stress, of flooding on victims in a post-flood context. |
| Mustaffa et al. | Values such as respect, sincerity, caring attitude, professionalism, and dignity maintenance are important in the communication discourse of community relief workers to help maintain victims' psychological health. |
| Shultz et al. | Implementing community-based strategies that reduce exposure to risk in the context of flooding can reduce the physical and psychological impact. The implementation of these strategies relies on resilience within the affected community. |
| Song & Li | Leadership, disaster awareness, perceptions of physical and social environments and well-being in social networks increase individual resilience. |
| Zhong et al. | Displaced persons to planned shelters following a flood have a lower prevalence of mental health problems such as anxiety, depression and post-traumatic stress disorder compared to non displaced persons. |

However, resilience is a multi-faceted topic, and social capital can be affected by the interactions between individuals and institutions or governments (Adger, 2001). This aspect is particularly important in the context of floods, as it has been shown that inadequate state or government responses can have a lasting impact in affected communities (Chioma et al., 2019; Tullos, 2018). In other words, resilience is not only determined by the interactions within a community, but can be improved (or damaged) by external factors. Because governments and authorities will continue to have a central role in the planning and implementation of adaptation measures to floods in the future, there is a need to determine how decisions by these entities influence the resilience of affected individuals and communities. So far, this topic remained unexplored in the studies analyzed, but it needs to be considered in future analyses that seek to provide a holistic understanding of resilience in the context of floods.

The studies in the second category analyzed the impact of external interventions on the mental health of flood victims, either from preventive measures (as in the case of Zhong et al. (2020) who analyzed the effect of planned sheltering in flood victims), or post-disaster measures (as in the case of Butler et al. (2018), who examined how decisions made by authorities in the aftermath of a flooding event impact the mental health of individuals or in the case of Mustaffa et al. (2018), who explored how the language used by relief workers reduced the mental health impact on flood victims). These studies assessed mental health outcomes that were different from resilience (e.g., anxiety, depression and post-traumatic stress disorder in the study of Zhong et al., and qualitative data about mental health and psychological well-being in general in the studies of Butler et al. and Mustaffa et al.).

Overall, these studies show that measures taken by authorities have an important impact on the mental health of those affected by floods. However, one important aspect that was not addressed in these studies is how measures taken within the community influence the mental health of its individuals. Usually, communities that have been affected by floods (or that are threatened by them) have some level of preparedness that is driven by actions at the local level, which can include certain intervention measures (such as preparing sandbags^23^, the implementation of early warning systems (Priyanti et al., 2019), or conducting risk reduction workshops (Shariff and Hamidi, 2019)). Sometimes, these measures can cause confusion or be dysfunctiona l(Khalid et al., 2015) when a disaster occurs, thereby negatively impacting the mental health of individuals. However, this aspect was not addressed in the analyzed studies in the second category but remains an important area that needs to be explored in order to determine the relative importance of external and internal interventions in a mental health context and to identify areas that need to be prioritized in the event of a disaster to ensure that individuals are able to be resilient.

Generally speaking, the studies analyzed in this scoping review presented results that were in agreement with other works in the area of climate change, resilience, and social capital (Carmen et al., 2022; Smith et al., 2012). However, certain limitations that affect the robustness of the results and that limit their application in a broader context were identified. First, in most of the studies, mental health information was provided by the victims themselves, according to their perception of their mental health status, which, at best, is only a rough estimate of the true mental health status of the individuals (North and Pfefferbaum, 2013). Ideally, the mental health status of an individual should be assessed by a mental health professional, and the fact that this was not considered in the analyzed studies raises the possibility of bias in the collected data, which would compromise the robustness of the findings.

Second, most of the studies utilized a cross-sectional design by collecting data after the flooding event, and because of this, there was no follow up over time to determine if the attitudes and beliefs from individuals (which were collected using a Likert scale in most cases) changed over time. The perceptions, attitudes, and responses from an individual are not stationary, and in the event of a flooding, it is important to quantify changes in these factors over time (Hudson et al., 2020) in order to not only establish relationships but to also adequately measure the dynamics of human behavior in this context (Bubeck et al., 2020). From all the studies analyzed, only the study of Butler et al. (2018) had a longitudinal design, but the qualitative nature of this study limited the possibility of obtaining numerical estimates of participants’ beliefs and attitudes and their change over time. Although performing longitudinal studies in flood victims is challenging due to inherent environmental and logistic limitations during this type of disaster (Hudson et al., 2020), there is a pressing need for studies that adhere to this methodology. Only by having a time-resolved view of mental health indicators it will be possible to measure not only if affected individuals have been able to be resilient, but also to determine which opinions and attitudes toward intervention measures change over time, which in turn can be used by decision makers to refine the implementation of such policies.

Finally, it should be noted that the small sample size or the sampling process used in the studies pose a significant limitation to the robustness of their findings. In the first case, the majority of the studies that followed a qualitative approach were based on a sample size that ranged between 9 to 13 individuals (Brockie and Miller, 2017; Butler et al., 2018; Mustaffa et al., 2018). Although the information provided by these individuals is valuable, the analysis in each paper was focused on communities or areas with populations orders of magnitude higher than these sample numbers, and therefore, it is likely that the findings in each case only represent a partial view of the effects of the flooding event analyzed. This is an important limitation as aspects derived from the experience of others victims, which could be equally important to contextualize the impact of floods on mental health might have been not identified. It is important to mention that the study of Shultz et al. also followed a qualitative approach, but relied on third-party information (census information, local data sources, and expert consultation) to create a profile of flood hazard for the communities analyzed. Because the sources of information used are not referenced, it is not easy to determine if the methodology used can be applied in a different context, considering that in other areas of the world there might be a limited amount or a complete lack of information sources that can be used to build a flood hazard profile of impacted communities.

In the second case, there were certain issues with the sampling processes used by studies that used a quantitative approach (using regression models to determine the association between different variables). These issues included the use of samples not representative of all affected areas (Ludin et al., 2019), a drastic reduction in the amount of observations due to dropping entries with missing data (Masson et al., 2019), low response rates (Babcicky and Seebauer, 2017; Ludin et al., 2019; Masson et al., 2019), not correcting for having low representativity of certain age groups (Zhong et al., 2020), and relying in self-assessment of resilience (Song and Li, 2019), which introduced subjective bias. Although these studies had significantly higher sampling numbers than the papers that used a qualitative approach, the problems with the sampling process raise the possibility that certain important factors were omitted, or deemed not significant in the analysis. Collecting data from from flood victims is a delicate task, but it is important that future studies that aim to quantitatively analyze the impact of intervention measures in flood victims are based on data that adequately reflects the socio-demographic characteristics of the affected population, and that minimizes the introduction of bias in order to ensure that the robustness of the findings.

There are some limitations to this scoping review. First, only two databases were considered for this review, and therefore, it is possible that a limited number of relevant studies indexed in other platforms might have been missed. Second, the research strategy was limited to studies written in English and French, which might have led to the omission of relevant studies written in languages specific to certain areas affected by floods (such as Spanish, Portuguese, etc.). Future studies will seek to broaden the choice of languages to incorporate studies that allow to make a geographically-based analysis. Finally, the number of studies analyzed is relatively small with heterogeneous methodologies, which complicates the ability to make generalizations that are broadly applicable. Nonetheless, the small number of studies analyzed in fact emphasizes the ongoing need of additional research at the intersection of mental health and natural disasters, which is likely to become more significant in the near future.

## 5. Conclusion

Flood frequency and severity has increased, and broader segments of the population are likely to be affected by them. Existing literature that examines the effects of adaptation measures on the mental health of flood victims, is limited, with the majority of the studies focused on the factors that affect resilience in flood victims. The applicability of the findings of these studies is limited due to small sample sizes, data representativity, and the introduction of biases in the analysis due to the sampling processes used.

Therefore, there is a pressing need for additional studies that examine how mental health in flood victims is affected by intervention measures as governments or public entities increase the use of these interventions to limit the effect of flooding events. It is necessary to develop public policies that adequately consider the impact of these measures in the mental health of flood victims in order to avoid unnecessary suffering in those most affected by these natural disasters.

## Data Availability

All data produced in the present study are available upon reasonable request to the authors.

## 5. List of abbreviations

MESH: Medical Subject Headings
PICO: Population, Intervention, Comparison, and Outcome
PRISMA: Preferred Reporting Items for Systematic Reviews and Meta-Analyses
FEM: Fatima El-Mousawi
RB: Rawda Berkat
BN: Bouchra Nasri
AM: Ariel Mundo Ortiz

## 7. Author Contributions

FEM and BN conceived and designed the study.

FEM, RB and BN collected data, performed data analysis and interpretation.

FEM, RB, BN and AM contributed to drafting the article.

FEM, RB, BN and AM contributed to the critical revision of the article.

FEB, RB, AM and BN contributed to final approval of the version to be submitted.

## References

Adger, W., 2001. Social capital and climate change.

Alderman, K., Turner, L.R., Tong, S., 2012. Floods and human health: A systematic review. Environ. Int. 47, 37–47. https://doi.org/10.1016/j.envint.2012.06.003

Allwood, J.M., Bosetti, V., Dubash, N.K., Gomez-Echeverri, L., von Stechow, C., 2014. Glossary., in: Climate Change 2014: Mitigation of Climate Change. Contribution of Working Group III to the Fifth Assessment Report of the Intergovernmental Panel on Climate Change. Cambridge University Press, Cambridge, United Kingdom and New York, NY, USA.

Amir-Behghadami, M., Janati, A., 2020. Population, Intervention, Comparison, Outcomes and Study (PICOS) design as a framework to formulate eligibility criteria in systematic reviews. Emerg. Med. J. https://doi.org/10.1136/emermed-2020-209567

Babcicky, P., Seebauer, S., 2017. The two faces of social capital in private flood mitigation: opposing effects on risk perception, self-efficacy and coping capacity. J. Risk Res. 20, 1017–1037. https://doi.org/10.1080/13669877.2016.1147489

Brink, E., Wamsler, C., 2019. Citizen engagement in climate adaptation surveyed: The role of values, worldviews, gender and place. J. Clean. Prod. 209, 1342–1353. https://doi.org/10.1016/j.jclepro.2018.10.164

Brockie, L., Miller, E., 2017. Understanding Older Adults’ Resilience During the Brisbane Floods: Social Capital, Life Experience, and Optimism. Disaster Med. Public Health Prep. 11, 72–79. https://doi.org/10.1017/dmp.2016.161

Bubeck, P., Berghäuser, L., Hudson, P., Thieken, A.H., 2020. Using Panel Data to Understand the Dynamics of Human Behavior in Response to Flooding. Risk Anal. 40, 2340–2359. https://doi.org/10.1111/risa.13548

Butler, C., Walker-Springett, K., Adger, W.N., 2018. Narratives of recovery after floods: Mental health, institutions, and intervention. Soc. Sci. Med. 216, 67–73. https://doi.org/10.1016/j.socscimed.2018.09.024

Carmen, E., Fazey, I., Ross, H., Bedinger, M., Smith, F.M., Prager, K., McClymont, K., Morrison, D., 2022. Building community resilience in a context of climate change: The role of social capital. Ambio 51, 1371–1387. https://doi.org/10.1007/s13280-021-01678-9

Chioma, O.C., Chitakira, M., Olanrewaju, O.O., Louw, E., 2019. Impacts of flood disasters in Nigeria: A critical evaluation of health implications and management. Jàmbá J. Disaster Risk Stud. 11. https://doi.org/10.4102/jamba.v11i1.557

Creswell, J.W., Poth, C.N., 2016. Qualitative Inquiry and Research Design: Choosing Among Five Approaches. SAGE Publications.

Doocy, S., Daniels, A., Murray, S., Kirsch, T.D., 2013. The Human Impact of Floods: a Historical Review of Events 1980-2009 and Systematic Literature Review. PLOS Curr. Disasters. https://doi.org/10.1371/currents.dis.f4deb457904936b07c09daa98ee8171a

Ebi, K.L., Semenza, J.C., 2008. Community-Based Adaptation to the Health Impacts of Climate Change. Am. J. Prev. Med., Theme Issue: Climate Change and the Health of the Public 35, 501–507. https://doi.org/10.1016/j.amepre.2008.08.018

Engle, N.L., de Bremond, A., Malone, E.L., Moss, R.H., 2014. Towards a resilience indicator framework for making climate-change adaptation decisions. Mitig. Adapt. Strateg. Glob. Change 19, 1295–1312. https://doi.org/10.1007/s11027-013-9475-x

Field, C.B., Barros, V., Stocker, T.F., Dahe, Q. (Eds.), 2012. Managing the Risks of Extreme Events and Disasters to Advance Climate Change Adaptation: Special Report of the Intergovernmental Panel on Climate Change, 1st ed. Cambridge University Press. https://doi.org/10.1017/CBO9781139177245

Herrman, H., Stewart, D.E., Diaz-Granados, N., Berger, E.L., Jackson, B., Yuen, T., 2011. What is Resilience? Can. J. Psychiatry 56.

Hudson, P., Thieken, A.H., Bubeck, P., 2020. The challenges of longitudinal surveys in the flood risk domain. J. Risk Res. 23, 642–663. https://doi.org/10.1080/13669877.2019.1617339

Jones, N., Clark, J.R.A., 2013. Social capital and climate change mitigation in coastal areas: A review of current debates and identification of future research directions. Ocean Coast. Manag. 80, 12–19. https://doi.org/10.1016/j.ocecoaman.2013.03.009

Khalid, M.S., Mustaffa, C.S., Marzuki, M.N., Sakdan, M.F., Sipon, S., Ariffin, M.T., Shafiai, S., 2015. Failure to React Positively to Flood Early Warning Systems: Lessons Learned by Flood Victims from Flash Flood Disasters: The Malaysia Experience 9.

Ludin, S.M., Rohaizat, M., Arbon, P., 2019. The association between social cohesion and community disaster resilience: A cross-sectional study. Health Soc. Care Community 27, 621–631. https://doi.org/10.1111/hsc.12674

Mann, M. (Michelle), Hosman, C.M.H., Schaalma, H.P., de Vries, N.K., 2004. Self-esteem in a broad-spectrum approach for mental health promotion. Health Educ. Res. 19, 357–372. https://doi.org/10.1093/her/cyg041

Mason, V., Andrews, H., Upton, D., 2010. The psychological impact of exposure to floods. Psychol. Health Med. 15, 61–73. https://doi.org/10.1080/13548500903483478

Masson, T., Bamberg, S., Stricker, M., Heidenreich, A., 2019. “We can help ourselves”: does community resilience buffer against the negative impact of flooding on mental health? Nat. Hazards Earth Syst. Sci. 19, 2371–2384. https://doi.org/10.5194/nhess-19-2371-2019

Milly, P.C.D., Wetherald, R.T., Dunne, K.A., Delworth, T.L., 2002. Increasing risk of great floods in a changing climate. Nature 415, 514–517. https://doi.org/10.1038/415514a

Mustaffa, C.S., Najib Ahmad Marzuki, Mohd Sukeri Khalid, Mohd Foad Sakdan, Sapora Sipon, 2018. Understanding Malaysian Malays communication characteristics in reducing psychological impact on flood victims. J. Komun. Malays. J. Commun. 34, 20–36.

Nofal, O.M., van de Lindt, J.W., 2020. High-resolution approach to quantify the impact of building-level flood risk mitigation and adaptation measures on flood losses at the community-level. Int. J. Disaster Risk Reduct. 51, 101903. https://doi.org/10.1016/j.ijdrr.2020.101903

North, C.S., Pfefferbaum, B., 2013. Mental Health Response to Community Disasters: A Systematic Review. JAMA 310, 507–518. https://doi.org/10.1001/jama.2013.107799

Osberghaus, D., Dannenberg, A., Mennel, T., Sturm, B., 2010. The Role of the Government in Adaptation to Climate Change. Environ. Plan. C Gov. Policy 28, 834–850. https://doi.org/10.1068/c09179j

Priyanti, R.P., Hidayah, N., Rosmaharani, S., Nahariani, P., Asri, Mukarromah, N., Mundakir, 2019. Community Preparedness in Flood Disaster: A Qualitative Study. Int. Q. Community Health. Educ. 40, 67–68. https://doi.org/10.1177/0272684X19853169

Richardson, G.R.A., 2010. Adapting to Climate Change: An introduction for Canadian Municipalities.

Saeed, S.A., Gargano, S.P., 2022. Natural disasters and mental health. Int. Rev. Psychiatry 34, 16–25. https://doi.org/10.1080/09540261.2022.2037524

Saulnier, D.D., Ribacke, K.B., Schreeb, J. von, 2017. No Calm After the Storm: A Systematic Review of Human Health Following Flood and Storm Disasters. Prehospital Disaster Med. 32, 568–579. https://doi.org/10.1017/S1049023X17006574

Sen, Z., 2018. Flood Modeling, Prediction and Mitigation. Springer International Publishing, Cham. https://doi.org/10.1007/978-3-319-52356-9

Shariff, N.N.M., Hamidi, Z.S., 2019. Community-based approach for a flood preparedness plan in Malaysia. Jàmbá J. Disaster Risk Stud. 11. https://doi.org/10.4102/jamba.v11i1.598

Sharifi, A., Pathak, M., Joshi, C., He, B.-J., 2021. A systematic review of the health co-benefits of urban climate change adaptation. Sustain. Cities Soc. 74, 103190. https://doi.org/10.1016/j.scs.2021.103190

Shultz, J.M., McLean, A., Herberman Mash, H.B., Rosen, A., Kelly, F., Solo-Gabriele, H.M., Youngs Jr, G.A., Jensen, J., Bernal, O., Neria, Y., 2013. Mitigating flood exposure. Disaster Health 1, 30–44. https://doi.org/10.4161/dish.23076

Shultz, J.M., Neria, Y., 2013. Trauma signature analysis. Disaster Health 1, 4–8. https://doi.org/10.4161/dish.24011

Smith, J.W., Anderson, D.H., Moore, R.L., 2012. Social Capital, Place Meanings, and Perceived Resilience to Climate Change*. Rural Sociol. 77, 380–407. https://doi.org/10.1111/j.1549-0831.2012.00082.x

Song, J., Li, W., 2019. Linkage Between the Environment and Individual Resilience to Urban Flooding: A Case Study of Shenzhen, China. Int. J. Environ. Res. Public. Health 16, 2559. https://doi.org/10.3390/ijerph16142559

Svetlana, D., Radovan, D., Ján, D., 2015. The Economic Impact of Floods and their Importance in Different Regions of the World with Emphasis on Europe. Procedia Econ. Finance, International Scientific Conference: Business Economics and Management (BEM2015) 34, 649–655. https://doi.org/10.1016/S2212-5671(15)01681-0

Tricco, A.C., Lillie, E., Zarin, W., O’Brien, K.K., Colquhoun, H., Levac, D., Moher, D., Peters, M.D.J., Horsley, T., Weeks, L., Hempel, S., Akl, E.A., Chang, C., McGowan, J., Stewart, L., Hartling, L., Aldcroft, A., Wilson, M.G., Garritty, C., Lewin, S., Godfrey, C.M., Macdonald, M.T., Langlois, E.V., Soares-Weiser, K., Moriarty, J., Clifford, T., Tunçalp, Ö., Straus, S.E., 2018. PRISMA Extension for Scoping Reviews (PRISMA-ScR): Checklist and Explanation. Ann. Intern. Med. 169, 467–473. https://doi.org/10.7326/M18-0850

Tullos, D., 2018. How to achieve better flood-risk governance in the United States. Proc. Natl. Acad. Sci. 115, 3731–3734. https://doi.org/10.1073/pnas.1722412115

Zhong, S., Pang, M., Ho, H.C., Jegasothy, E., Clayton, S., Wang, Z., Huang, C., 2020. Assessing the effectiveness and pathways of planned shelters in protecting mental health of flood victims in China. Environ. Res. Lett. 15, 125006. https://doi.org/10.1088/1748-9326/abc446

Zhou, H., Wang, J., Wan, J., Jia, H., 2010. Resilience to natural hazards: a geographic perspective. Nat. Hazards 53, 21–41. https://doi.org/10.1007/s11069-009-9407-y

