## Supplement material for "The Impact of Flood Adaptation Measures on Affected Population’s Mental Health: A mixed method Scoping Review"

**Table 1 – Medline search strategy**

| No. | Queries |
| --- | --- |
| 1 | Disaster Planning/ |
| 2 | Floods/ or Disasters/ or Rivers/ or Lakes/ or Rain/ or Meteorological Concepts/ |
| 3 | (Inundation* or Flood* or Heavy precipitation* or rain or rains or river* or lake* or high water).ab,kw,ti. |
| 4 | 2 or 3 |
| 5 | Mental Health/ or Adaptation, Psychological/ or Psychological Distress/ or Psychological Trauma/ or Resilience, Psychological/ or Substance-Related Disorders/ or Mental Disorders/ |
| 6 | exp Mental Disorders/ |
| 7 | 5 or 6 |
| 8 | (Mental health or Psychological or Substance abuse or Mental disorder* or Mental illness* or Distress).ab,kw,ti. |
| 9 | 7 or 8 |
| 10 | (Disaster* adj2 (Risk* or reduction or preparedness or resilience or planning or management)).ab,kw,ti. |
| 11 | (Adaptation adj2 (planning or strateg* or measure* or decision* or approach*)).ab,kw,ti. |
| 12 | 1 or 10 or 11 |
| 13 | 4 and 9 and 12 |

**Table 2 – Web of Science search strategy**

| No. | Queries |
| --- | --- |
| 1 | TS=(Disaster* NEAR/2 (Risk* or reduction or preparedness or resilience or planning or management)) (Exclude – Database) |
| 2 | TS=(Adaptation NEAR/2 (planning or strateg* or measure* or decision* or approach*)) (Exclude – Database) |
| 3 | #2 OR #1 (Exclude – Database) |
| 4 | TS=(Inundation* or Flood* or “Heavy precipitation” or rain or rains or river* or lake* or “high water") (Exclude – Database) |
| 5 | TS=("Mental health” or Psychological or “Substance abuse” or “Mental disorder” or “Mental illness” or Distress) (Exclude – Database) |
| 6 | #3 AND #4 AND #5 (Exclude – Database) |
